# *Real-World Costs and Cost-Effectiveness of Next-Generation Insecticide Treated Bednets:* controlled interrupted time-series based economic evaluations of dual–active ingredient (AI) or piperonyl butoxide (PBO) insecticide-treated bed nets (ITNs) versus pyrethroid-only ITNs for malaria prevention

**DOI:** 10.64898/2026.06.18.26355830

**Authors:** Joshua O. Yukich, Peder Digre, Joseph Wagman, Eli Santiago, Maya Schane, Adama Gansané, Baltazar Candrinho, Christen Fornadel, Molly Robertson

**Affiliations:** Celia Scott Weatherhead School of Public Health and Tropical Medicine, Tulane University; PATH; Institut National de Sante Publique; Ministerio da Saude; Innovative Vector Control Consortium; The Global Fund

## Abstract

Growing insecticide resistance and other causes of residual transmission of malaria parasites have led to a global stagnation in progress in malaria control, despite widespread deployment of insecticide-treated bed nets (ITNs). New bed net technologies including dual active ingredient (AI) and piperonyl butoxide plus pyrethroid (PBO + P) nets have been developed to address these problems and be more effective malaria prevention tools. Two types of dual-AI ITNs and PBO + P ITNs were evaluated across five countries in pilot studies coupled with economic evaluations. Chlorfenapyr + pyrethroid (C + P) ITNs showed the best overall performance in terms of incidence rate reduction (Incidence rate ratio 0.62 vs. standard ITN) and generally in terms of incremental cost-effectiveness ratio, but using any dual-AI or PBO + P ITN was favored over standard pyrethroid-only ITNs alone. There was significant overlap between the recommended strategy in any given study location due to high uncertainty in cases averted and relatively similar product costs. While the addition of indoor residual spraying (IRS) to standard ITNs was a more effective strategy than dual-AI or PBO + P ITNs, this evidence was based on only one study site and the incremental cost for the addition of IRS was substantially greater than the incremental cost of dual-AI or PBO ITNs compared to standard ITNs (ICER 18.59 USD per case averted vs. 1.51 USD per case averted for C + P ITN in this settings). The switch from standard ITNs to dual-AI or PBO ITNs is recommended in most African settings, with the chlorfenapyr + pyrethroid (C + P) nets being the broadly favored choice. Ultimately programs will need to incorporate local information on the malaria burden and up to date product price information to determine the most efficient malaria prevention strategy in the settings in which they serve.

## BACKGROUND

Since the beginning of the widespread deployment of insecticide-treated bed nets (ITN) to prevent malaria in Africa the malaria burden has been reduced dramatically (1). The global reduction in burden has been largely attributed to widespread distribution and use of pyrethroid-based ITNs (1). In recent years the decline in burden has stalled and malaria burden remains high and persistent in much of sub-Saharan Africa (2). Reasons for the stagnation are likely myriad and include stagnant funding levels, which are likely to be exacerbated by recent dramatic cuts in United States foreign assistance for health (3).

In addition, widespread pyrethroid resistance may be reducing the effectiveness of standard insecticide-treated bed nets (ITNs) in the prevention of malaria though the extent of the impact is uncertain and debated (4–9). Several types of ITNs that address pyrethroid resistance have been developed (10–18).

Optimal allocation of the multiple ITN types requires data on their performance in varied transmission and resistance contexts combined with information on the cost of deployment, especially in the context of reduced funding for malaria control.

The New Nets Project (NNP) was developed as an evidence generation and market shaping intervention to help expand market access for new ITN types while also generating evidence of their effectiveness and cost to better guide policy development around these new products. The NNP covered several types of ITN products including: 1) standard pyrethroid-only ITNs, 2) piperonyl-butoxide (PBO) (a pyrethroid synergist) + pyrethroid (PBO+P) ITNs; 3) pyriproxyfen (a juvenile growth inhibitor) + pyrethroid (Py+P) ITNs, and 4) chlorfenapyr (a novel AI insecticide) + pyrethroid (C+P) ITNs (19).

While standard ITNs are well documented to be cost-effective interventions, the relative cost-effectiveness of the addition of a second AI in a variety of transmission and resistance contexts is less well studied (9,20–24). In addition, robust cluster randomized controlled trials have shown that dual-AI ITNs containing chlorfenapyr or ITNs containing PBO have improved effectiveness compared to standard ITNs in some settings and that the addition of pyriproxyfen to a pyrethroid may also improve their effectiveness in some settings (13,14,16,17,25,26).

While these trials have demonstrated the superiority in effectiveness and cost-effectiveness of dual-AI or PBO ITNs in some settings (14,26,27), trial data can suffer from an inability to predict the gap in efficacy to effectiveness both from an epidemiological perspective and a cost and operational perspective. Data from controlled trials also suffers from a lack of generalizability to settings where trials were not conducted (28). Real-world data on effectiveness and cost can be useful to supplement trial data on costs and epidemiological effects to quantify the effectiveness–efficacy gap and to estimate real-world cost-effectiveness. Further, such data can also be used to validate the predictions of models derived from trial data.

Standard ITNs have a lower procurement cost than the dual-AI or PBO ITNs; as such, the marginal improvement in effectiveness needed to recommend switching must be determined in a variety of different settings. Further, the implications of switching to novel product types on distribution systems and their costs have not been well documented.

This study details the effectiveness, cost, and cost-effectiveness results of a series of observational (quasi-experimental) studies and costing studies to establish the incremental effectiveness and the incremental cost-effectiveness of dual-AI or PBO ITNs compared to standard ITNs in real-world settings.

## METHODS

### Description of observational studies

Five observational studies of the effectiveness of dual-AI or PBO ITNs were conducted in four study locations: Burkina Faso, Mozambique (two locations: North and West), Nigeria, and Rwanda. In each of the effectiveness study locations, dual-AI or PBO ITNs were distributed as part of mass campaigns in some preselected study areas (districts or local government areas) and standard ITNs were distributed in nearby study areas. In one study area in Rwanda, indoor residual spraying (IRS) was conducted three times over the study period in addition to the distribution of standard ITNs. The use of dual-AI or PBO ITNs or IRS plus standard ITNs comprise the *intervention* compared to standard ITNs alone. All effectiveness studies followed a controlled interrupted time series design. Details on the exact composition of each study are presented in Table 1 and Figure 1. Details of the methods of the studies as well as baseline prevalence, the primary vector species, and pyrethroid resistance status in each study area are reported elsewhere (19). In addition to the collection of data in the five effectiveness locations, cost data was collected based on nationwide costing exercises for distribution of standard and dual-AI or PBO ITNs in Mali and Uganda, while effectiveness data were not collected in these locations as part of the New Nets Project.

**Figure 1:**
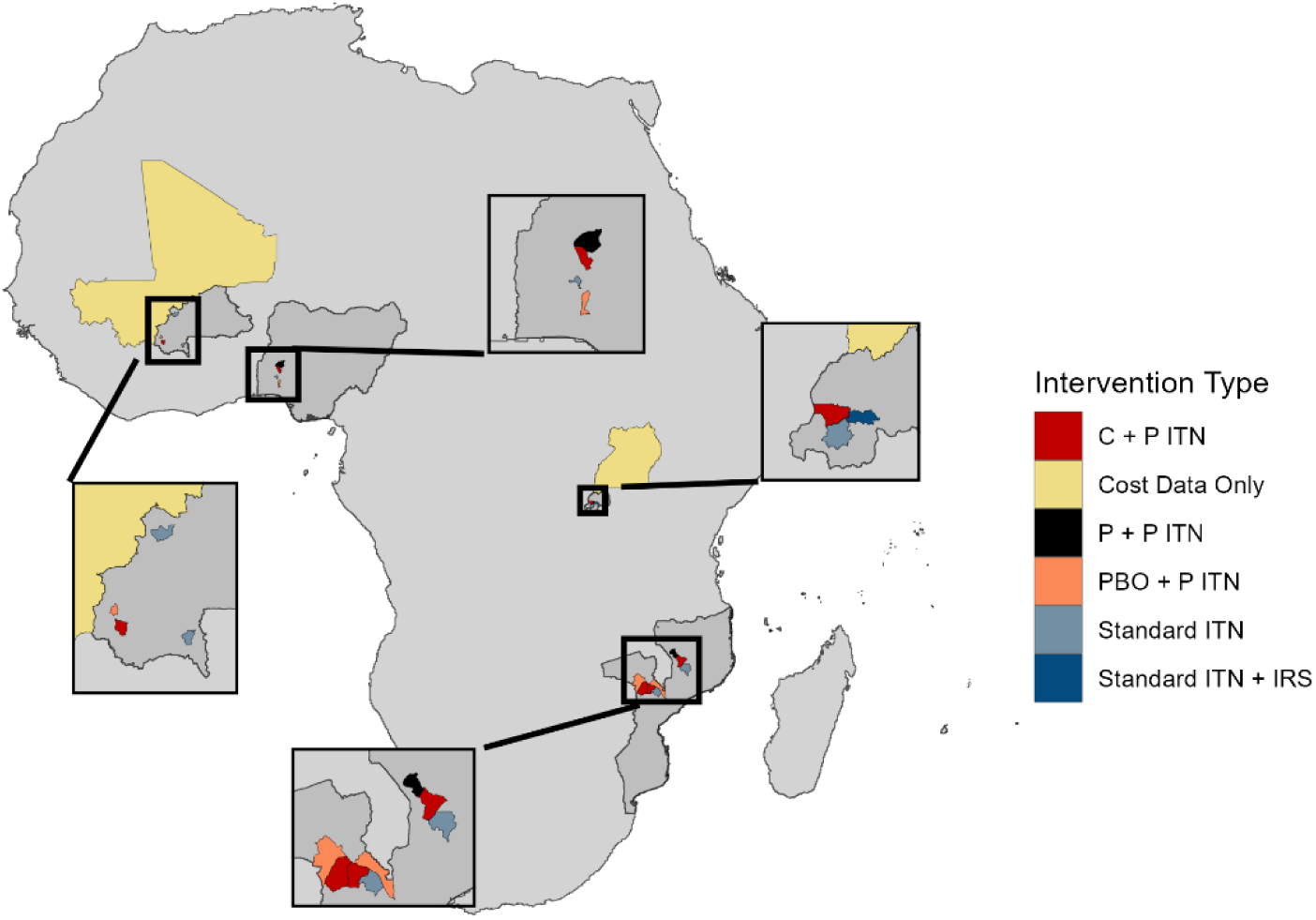
Study Locations, interventions and data collections included across all planned study sites (Mali, Mozambique, Burkina Faso, Uganda, Rwanda, Nigeria).

**Table 1.**
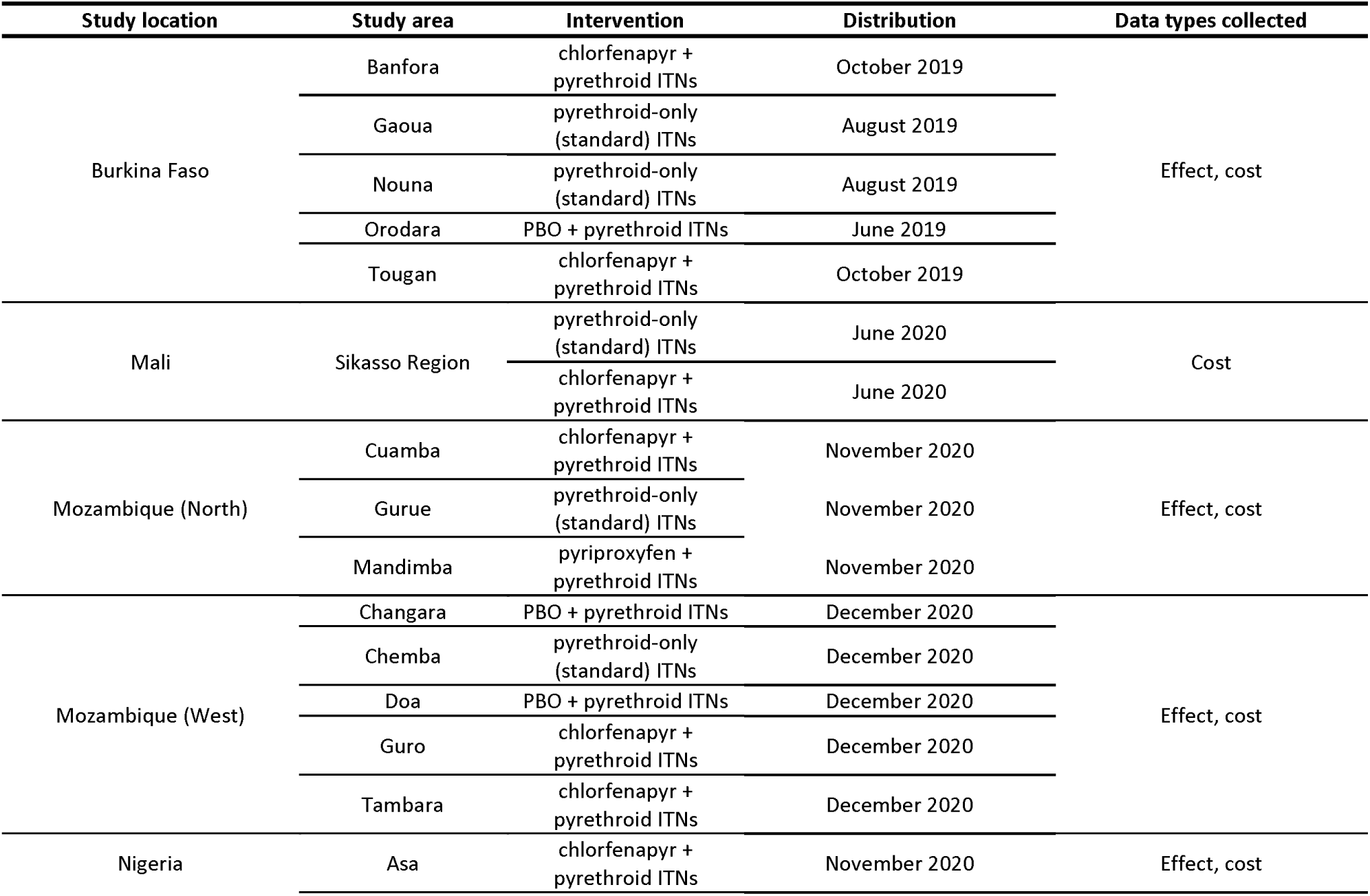
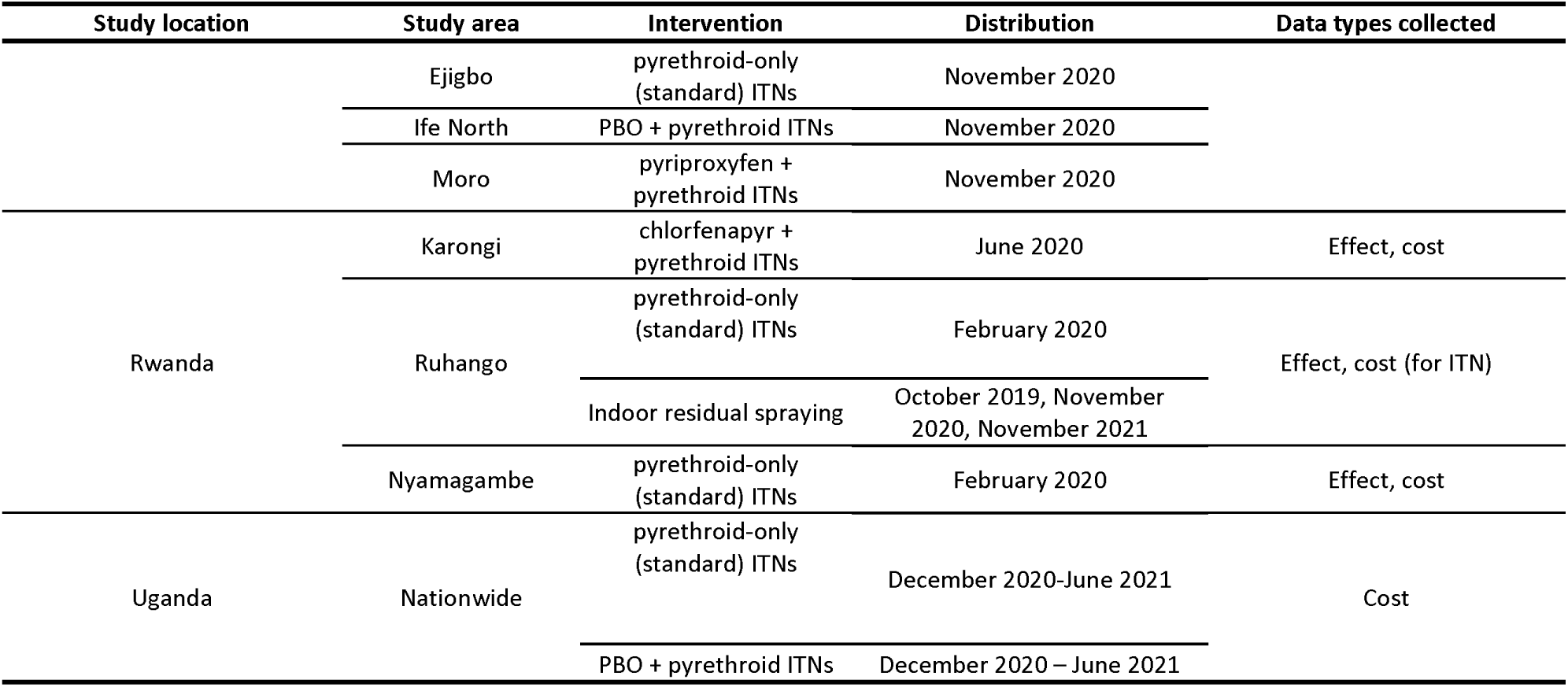
Description of operational pilot studies and real-world cost data sources.

For all observational studies, standard ITNs and dual-AI or PBO ITNs were delivered nearly simultaneously as part of mass campaigns, with different ITN types assigned by study area. Outcomes were monitored using cross-sectional surveys of parasite prevalence as well as monitoring of clinical malaria episodes through the routine health system (passive case detection). In each study area, data on cases of clinical malaria (confirmed using parasitological testing) was tracked for a minimum of twelve months prior to the intervention and twenty-four months post intervention.

### Effectiveness analysis

The number of cases of clinical malaria averted in each study area was estimated by fitting statistical models to the clinical malaria datasets. Models followed an interrupted time-series formulation and utilized a negative binomial likelihood. All models were fit in R version 4.5.1 (29) using the MASS or lme4 packages (30,31). Estimates of the average treatment effect were made for each year post-deployment of ITNs. Separate estimates were made for each study location, intervention, and year post-deployment. Statistical models were used to project the impact of intervention in each study area in observed scenarios as well as in counter-factual scenarios in which it was assumed that standard ITNs had been distributed in those areas as opposed to the intervention which had, in reality, been distributed. Cases averted were calculated by subtracting the number of cases projected from the model from the number of cases projected in the counterfactual (standard ITN scenario). Uncertainty around the cases averted estimates was calculated by making 10,000 simulated datasets in observed scenarios and counterfactual scenarios from the study sites specific models and calculating cases averted for each of these new datasets. Ninety-five percent credibility bounds for the cases averted estimates were then created from the 2.5% and 97.5% quantiles of the resulting cases averted distribution.

In addition to study-specific models, estimates of average treatment effect were also included in a pooled meta-analysis by taking study-specific effect estimates of the expected incidence rate ratio for each year post-deployment and their associated uncertainty (standard error) estimates as individual data points (studies). The meta-analysis followed a Der-Simonon-Laird random effects model formulation and was conducted using the *metafor* package (32).

Cases averted were converted to disability-adjusted life years (DALYs) averted using a simple linear approach in which the number of all cases averted used to calculate an estimated number of deaths averted assuming a case-fatality rate of 0.06%. Cases and deaths averted were then converted to DALYs averted assuming that each case averted was equivalent to 0.02 DALYs averted, and each death averted equivalent to 33 DALYs averted.

### Cost collection and analysis

The costs of distribution for standard ITNs and dual-AI or PBO ITNs through mass campaigns were established using a standardized approach to the estimation of program costs which was employed across all study sites. A description of each program/intervention was developed based on document reviews (operational records and logistics documents from key stakeholders, donors, and partner organizations), and key informant interviews. The intervention descriptions guided all further cost data collection.

The cost analyses followed a micro-costing or ingredients approach, where costs were estimated by first identifying all resources needed to deploy the interventions, secondly quantifying the amounts of those resources required and finally, establishing prices for all input resources.

Cost data were collected retrospectively from financial and operational records, reports, pay scales, budgets, fixed assets registries, and invoices as well as operational protocols and reports. Stakeholder interviews were utilized to capture costs or resource inputs not reflected in financial or other records, such as volunteer time or assets used during the distribution but owned prior to the distribution (i.e., vehicles), and national, regional, and local level supervision time, or shared resources that are not included in standard financial reports.

Key informants were identified through collaboration with national malaria program leadership and interviewed. Key informants were ideally chosen to be individuals heavily involved with the financing/accounting or planning/logistics within the organizations responsible for ITN distribution. Identification, collection, and review of documents such as financial and operational records, reports, pay scales, budgets, and invoices as well as operational protocols and reports accompanied these interviews. Interviews with all key informants were conducted remotely due to the COVID-19 pandemic.

In general, the time horizon for cost analyses was three years, corresponding to an assumed three-year lifetime of an ITN. Costs were divided into capital and recurrent costs, based on the lifetime of the goods or service being purchased. Capital costs were discounted in economic analysis using lifetimes determined through stakeholder interviews, expert information, and prior published literature at a discount rate of 3%. Varying discount rates and lifetimes were examined in a sensitivity analysis. Costs were also categorized by line-item group into the following categories: ITNs, co-payments, logistics, meeting costs, other direct costs, personnel, and procurement. The co-payments were payments made to manufacturers of ITNs by donor agencies (e.g. the Global Fund through the New Nets Project) to reduce the cost of dual-AI or PBO ITNs that was experienced by the individual country procurers. The ITN category represents amounts paid for the ITN commodities directly by countries (typically with support of the US President’s Malaria Initiative (PMI) or the Global Fund). These costs, as well as co-pay costs are treated as capital expenditure in the analysis.

All costs were converted to a common year and currency, to ensure comparability between locations with different inflation rates and currency values (2020 USD). Costs were converted from local currency to USD using the exchange rate in the year the cost analyses are conducted (33). The US consumer price index (CPI) was used to inflate or deflate all costs to 2020 USD (34). Total program costs were paired with the number of ITNs distributed to estimate the total cost per ITN distributed in each study area. Incremental costs were calculated based on the difference between the estimated cost of distributing the intervention in the study area and the estimated amount of conducting a standard ITN distribution in the same study area.

Costs for IRS were derived from prior real-word review of IRS cost data (35). Costs of treatment were derived from a systematic review of the literature [*in preparation*]. These results were disaggregated by treatment for uncomplicated vs. severe malaria and were considered only from the provider’s perspective. Mean and median costs as well as the distribution of costs were considered, in the review but only costs for the treatment of uncomplicated malaria are used in this synthesis. Costs of treatment were considered to linearly offset costs of prevention when net costs were calculated.

### Estimation of cost-effectiveness

The incremental cost-effectiveness of the use of each dual-AI or PBO ITN as compared to a standard ITN was estimated in each study area where that dual-AI or PBO ITN was deployed and is reported in terms of the incremental cost-effectiveness ratio. The ratio is calculated using the following formula:

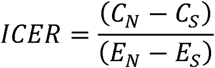

where *C_N_* is the cost of the dual-AI or PBO ITN and *C_S_* is the cost of the standard ITN and *E_N_* is the effect of the dual-AI or PBO ITN in terms of clinical cases averted and *E_S_* is the effect of the standard ITN in terms of clinical cases averted. Because the standard ITN is considered the reference scenario and cases averted are calculated relative to this scenario the formula reduces to:

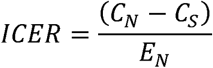

where the cases averted are calculated per the methods outlined above. In the case of gross ICER the costs reflect the cost above, not including any cost savings from cases averted. In the net cost scenarios, the ICER will reflect the following modification:

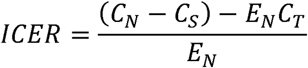

where *C_T_* is the cost of treatment and thus the ICER reflects the net cost of averting cases from the use of a dual-AI or PBO ITN compared to the use of standard ITNs. Cost savings are calculated assuming that clinical cases averted result in saving of the cost of treatment for each averted case. Costs of treatment were simulated for each case averted simulation using a gamma distribution with a shape and scale determined based on the cost of treatment systematic review described above (shape = 7.60, scale = 1).

In Rwanda, one study area deployed a combination of IRS and standard ITNs. In this case, the cost of IRS is treated identically to the use to dual-AI or PBO ITNs and the incremental cost-effectiveness ratio reflects the use of IRS and standard ITNs compared to standard ITNs alone.

Cost-effectiveness results are summarized using visualization and measures of central tendency and dispersion, as well as cost-effectiveness acceptability curves. The range of willingness-to-pay thresholds considered include from zero to at least three times gross domestic product (GDP) per capita for each target country to ensure that a full range of probable thresholds are considered.

### Sensitivity analyses

Several sensitivity analysis approaches were used in this study. One-way sensitivity analyses and scenario analyses were used in the analysis of cost data from each campaign. Though results of these analyses are not reported here, these analyses involved varying specific parameters one at a time and or in comparison to determine the impact of underlying assumptions such as discount rate and net price on the overall cost estimates. For cost-effectiveness analyses, probabilistic sensitivity analyses were conducted using Monte Carlo simulation considering uncertainty in effectiveness and cost arising from each scenario. Additionally, a global sensitivity analysis was conducted to integrate information across study locations and years while still considering uncertainty in effect and cost. In the global sensitivity analysis uncertainty in effect was derived from meta-analytic models and uncertainty in cost was derived from synthesis of cost data across all study locations by assuming that cost of delivery per ITN would follow a gamma distribution with shape and scale derived from individual study estimates (shape = 0.60, scale = 1). Monte Carlo simulation was then used to estimate the incremental cost-effectiveness of the use of dual-AI or PBO ITNs (or addition of IRS) compared to standard ITNs alone over a range of baseline malaria incidences with standard ITN programs in place.

### Budget impact

The total cost of deployment of dual-AI or PBO ITNs was calculated and reported as total and incremental estimates (the expenditure in excess of a standard ITN program) over a three-year time-period. Additionally, the cost of treatment of an uncomplicated malaria case was assumed based on a review of published literature [*in preparation*]. Budget impact is reported thus as gross impact (*i.e*. assuming no treatment cost savings) and as net impact (i.e. assuming full recovery of the cost of treatment for each case averted (or increase in budget required where cases increased).

## RESULTS

### Effectiveness

Routine data from each site was collected for a pre- and post-deployment period of a minimum of one year and up to three years in each site; data from Nigeria derived from the routine system was of poor quality and is not included further for effectiveness. The incidence of clinical malaria cases identified through the routine systems in each remaining study site are shown in Figure 2 below.

**Figure 2:**
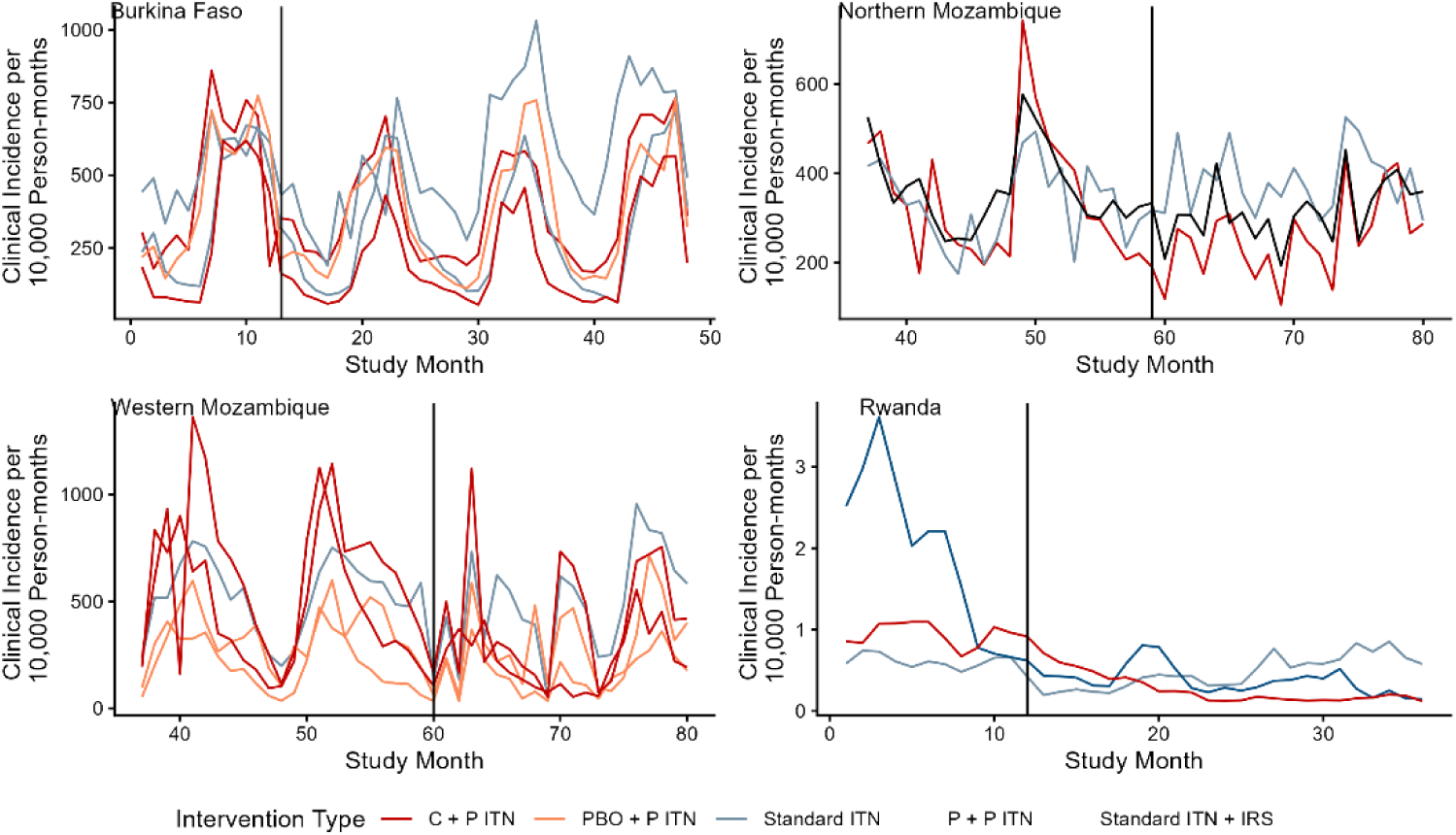
Incidence of clinical malaria in study areas.

For all study areas, negative binomial regression models were successfully fit, including interaction terms between year post-deployment of the intervention and the intervention type/area. The coefficient estimates from these terms are estimates of the average treatment effect of the incremental effect of the use of dual-AI or PBO ITNs (or IRS + standard ITNs) compared to standard ITNs. Model estimates are shown in Table 2 below.

**Table 2:** Average treatment effect estimates for each dual-AI or PBO ITN type (or IRS + standard ITNs) compared to standard ITNs.

| Interaction term | Burkina Faso | Mozambique (North) | Mozambique (West) | Rwanda |
| --- | --- | --- | --- | --- |
| PBO + pyrethroid ITN (Year 1) | 0.96<br>(1.29-0.71) |  | 0.98<br>(1.48-0.64) |  |
| PBO + pyrethroid ITN (Year 2) | 0.79<br>(1.07-0.59) |  | 0.88<br>(1.38-0.55) |  |
| PBO + pyrethroid ITN (Year 3) | 0.78<br>(1.05-0.58) |  |  |  |
| chlorfenapyr + pyrethroid ITN (Year 1) | 0.93<br>(1.19-0.73) | 0.56<br>(0.7-0.45)*** | 0.7<br>(1.05-0.46) | 0.69<br>(0.99-0.48)* |
| chlorfenapyr + pyrethroid ITN (Year 2) | 0.79<br>(1.01-0.62) | 0.75<br>(0.95-0.60)* | 0.57<br>(0.9-0.36)* | 0.15<br>(0.21-0.10)*** |
| chlorfenapyr + pyrethroid ITN (Year 3) | 0.86<br>(1.09-0.67) |  |  |  |
| pyriproxyfen + pyrethroid ITN (Year 1) |  | 0.7<br>(0.87-0.56)** |  |  |
| pyriproxyfen + pyrethroid ITN (Year 2) |  | 0.82<br>(1.03-0.65) |  |  |
| IRS (Year 1) |  |  |  | 0.46<br>(0.65-0.32)*** |
| IRS (Year 2) |  |  |  | 0.15<br>(0.22-0.11)*** |
| $p < 0.05$, ** $p < 0.01$, *** $p < 0.001$ | | | | |

PBO + pyrethroid ITNs were always seen to be protective relative to standard ITNs but in no case was the individual effect estimate significant at the 5% level. Chlorfenapyr + pyrethroid ITNs were seen to be protective. Pyriproxyfen + pyrethroid ITNs were also seen to be protective relative to standard ITNs in in northern Mozambique (the only location where deployment occurred with useable data) and IRS + standard ITNs was also highly protective relative to standard ITNs alone in Rwanda (also the only location where this intervention was deployed).

Based on meta-analysis of the individual study area and year results, the interventions deployed reduced malaria incidence relative to the deployment of standard ITNs with an average effect corresponding to a nearly 30 percent reduction in the incidence of clinical malaria detected in the health systems (Figure 3). The largest effects were seen in Rwanda where the introduction of chlorfenapyr + pyrethroid ITNs or IRS + standard ITNs reduced malaria incidence by more than 50%. The effect of chlorfenapyr + pyrethroid ITNs were much higher (corresponding to a nearly 40% reduction in malaria incidence (55% - 14%)). Pyriproxyfen + pyrethroid ITNs were considered effective across study sites in two countries with an effect corresponding to an approximately 25% reduction in malaria incidence (36% - 12%). While PBO + pyrethroid nets were less effective, with an average effect size of only around 14% (26% - 0%) reduction in malaria relative to standard ITNs Regression models were used to simulate interventions data sets under the as observed and in counter factual (as if standard ITNs had been deployed in the area) datasets in order to estimate and quantify the uncertainty around estimates of cases averted (Figure 4).

**Figure 3.**
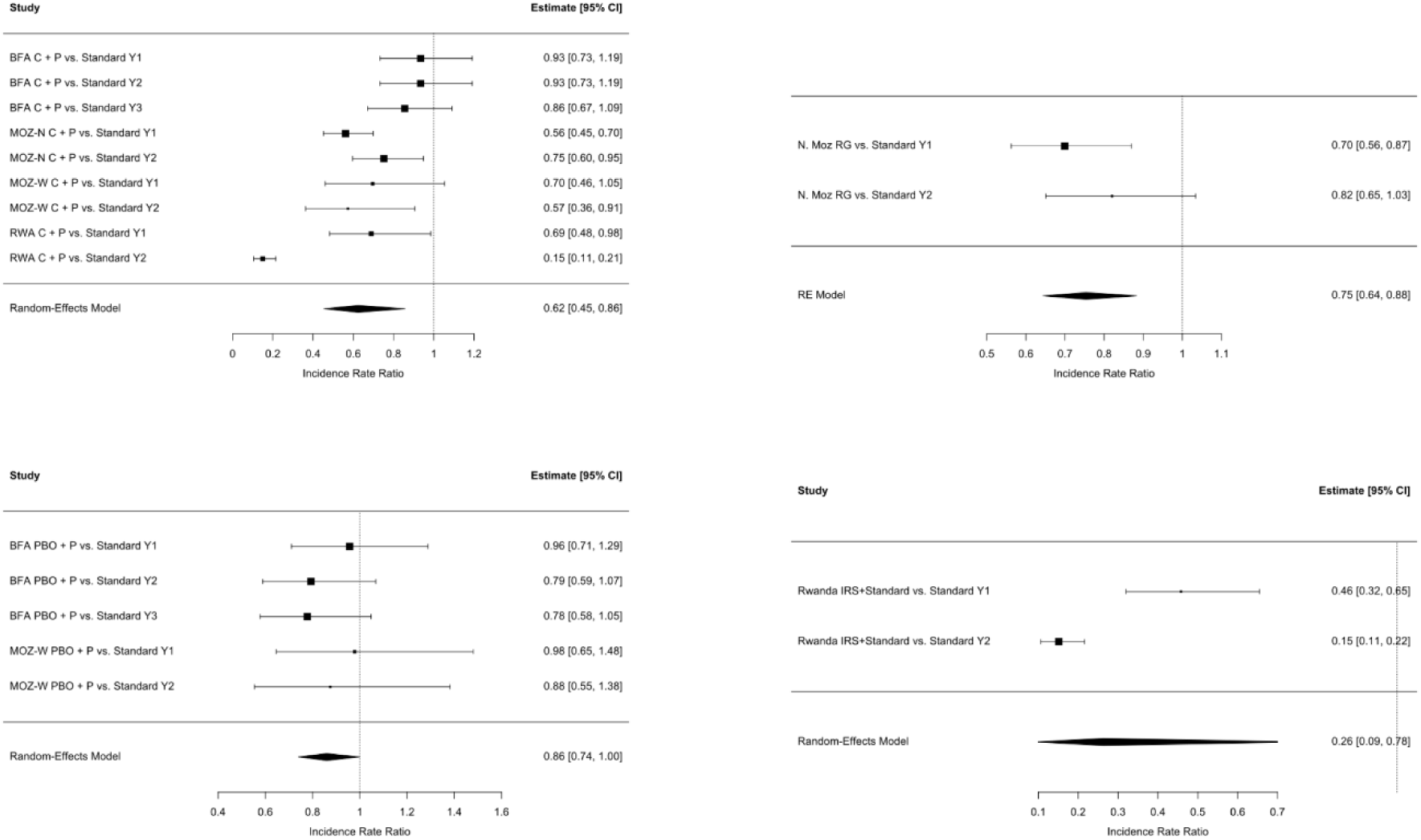
Caterpillar plot results of meta-analysis of annual effect estimates of dual-AI or PBO ITNs (or IRS + standard ITNs) compared to standard ITNs alone.

**Figure 4:**
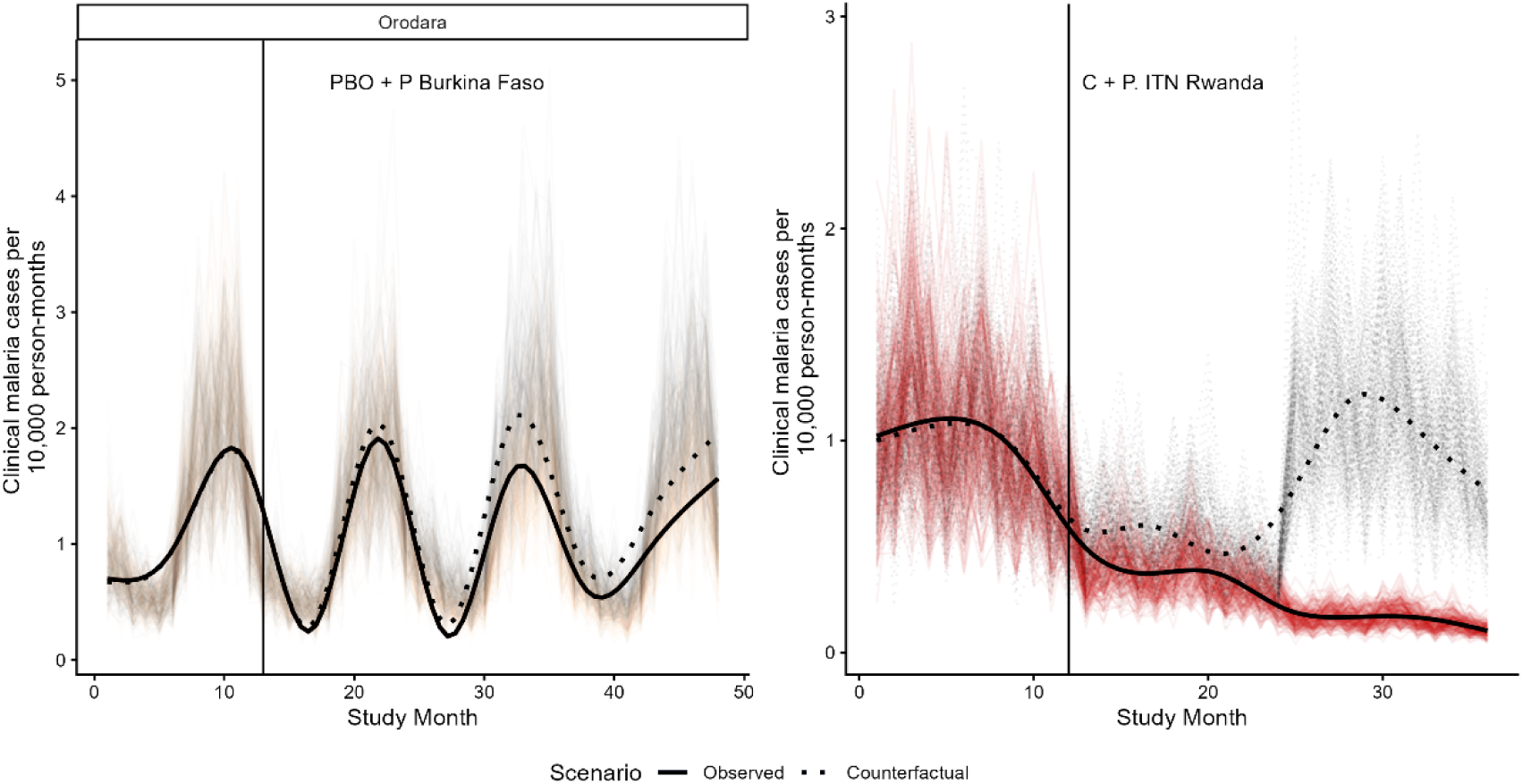
Examples of observed and counter-factual simulated results for PBO + pyrethroid ITNs vs. standard LLIN in Orodara District, Burkina Faso and chlorfenapyr + pyrethroid ITNs vs. standard ITNs in Rwanda. Counter-factual simulations are shown in grey.

Cases averted per month and cumulatively over the study periods were calculated as the difference between each pair of simulations (as observed and counter factual) (Figure 5) and are shown with credibility bounds in Table 3 and visually in Figure 5.

**Figure 5:**
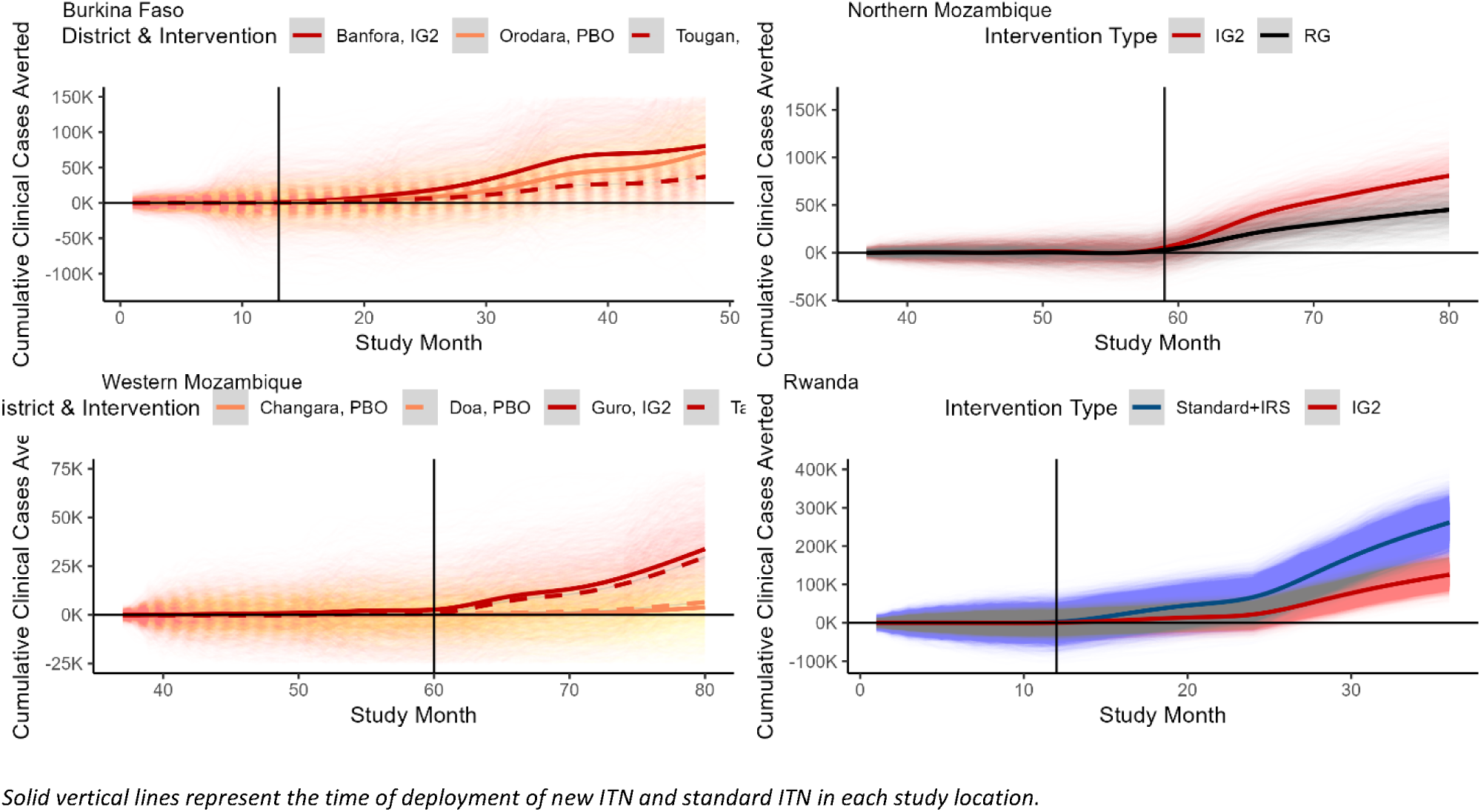
Cumulative cases averted relative to standard ITN deployment in study sites.

**Table 3:** Cases averted during the study period by intervention.

| Study location | Study area | Intervention | Cumulative cases averted | Study period (years) |
| --- | --- | --- | --- | --- |
| Burkina Faso | Banfora | chlorfenapyr + pyrethroid ITN | 103,662<br>(-24,707 – 230,164) | 3 |
|  | Tougan | chlorfenapyr + pyrethroid ITN | 35,804<br>(-7,249 – 84,634) |  |
|  | Orodara | PBO + pyrethroid ITN | 71,030<br>(-878 – 141,018) |  |
| Mozambique (North) | Cuamba | chlorfenapyr + pyrethroid ITN | 79,551<br>(44,987 – 116,554) | 2 |
|  | Mandimba | pyriproxyfen + pyrethroid ITN | 44,614<br>(15,679 – 72,694) |  |
| Mozambique (West) | Guro | chlorfenapyr + pyrethroid ITN | 33,543<br>(-5,350 – 71,620) | 2 |
|  | Tambara | chlorfenapyr + pyrethroid ITN | 29,460<br>(-7,494 – 65,092) |  |
|  | Changara | PBO + pyrethroid ITN | 3,769<br>(-18,534 – 26,537) |  |
|  | Doa | PBO + pyrethroid ITN | 4,913<br>(-23,581 – 34,900) |  |
| Rwanda | Karongi | chlorfenapyr + pyrethroid ITN | 124,212<br>(84,956 – 164,673) | 2 |
|  | Ruhango | IRS + standard ITN | 260,188<br>(187,154 – 334,717) |  |

The expected cases averted varied depending on the study location and type of ITN, with higher baseline malaria incidence leading to higher numbers of cases averted, and the addition of IRS to standard ITNs averted more cases than other interventions, but with chlorfenapyr + pyrethroid ITNs generally next, followed by pyriproxyfen + pyrethroid ITNs and finally PBO + pyrethroid ITNs.

### Costs

The costs of ITN distribution were evaluated across pilot observational study locations as well as in two additional study sites where dual-AI or PBO ITNs had been distributed. Intervention descriptions generally found that no major changes to program design or implementation were necessary for the distribution of dual-AI or PBO ITNs as compared to standard ITNs. Unit cost estimates and line-item cost breakdowns are shown in Figure 6.

**Figure 6:**
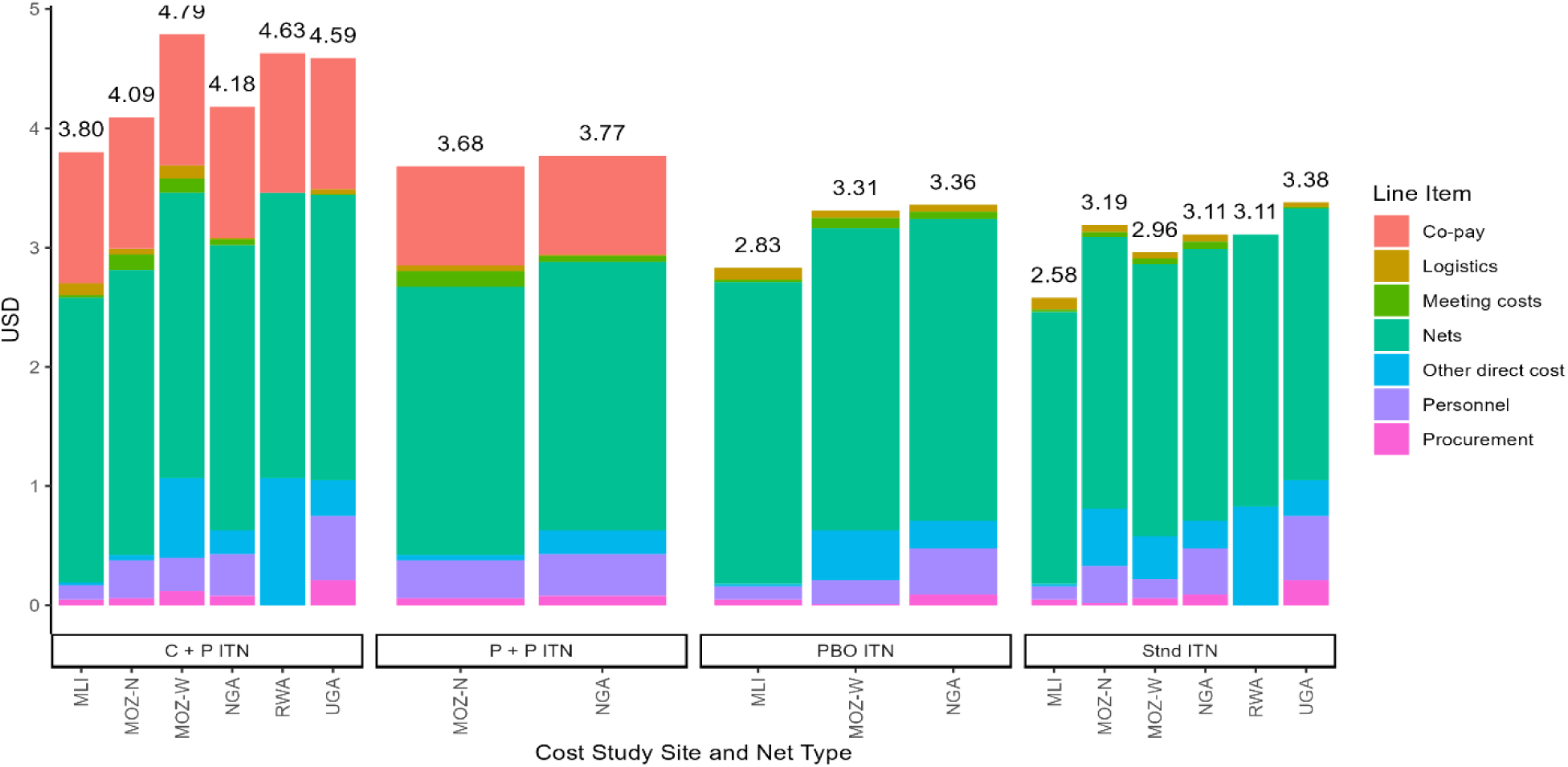
Costs per ITN distributed (USD).

Costs per ITN distributed varied between locations and net types but variation in total costs per net distributed were driven mainly by the co-pay (equivalent to the excess cost of new ITNs compared to standard ITN). When co-pays were excluded, costs per ITN distributed were similar across all ITN types and study locations. Costs per ITN distributed ranged from approximately 3 USD to 5 USD. The ITNs themselves accounted for the largest share of all costs with non-ITN (distribution costs) estimated to represent approximately 15-30% of total costs of distribution and the ITNs to represent 70-85% of the total.

### Cost per case averted

Costs per ITN distributed can be combined with estimates of cases averted in each study area to produce estimates of cost per case averted by ITN type in each study location. Results are summarized in Table 4.

**Table 4:** Cost per Case Averted Estimated During the New Nets Project.

| Study location | Study area | Intervention | Cumulative cases averted | Population | Total incremental cost (USD) | Incremental cost per case averted (USD) |
| --- | --- | --- | --- | --- | --- | --- |
| Burkina Faso | Banfora | chlorfenapyr + pyrethroid ITN | 103,662 | 423,519 | 287,052 | 2.77 |
|  | Tougan | chlorfenapyr + pyrethroid ITN | 35,804 | 310,919 | 210,734 | 5.89 |
|  | Orodara | PBO +<br>pyrethroid<br>ITN | 71,030 | 257,258 | 35,730 | 0.50 |
| Mozambique<br>(North) | Cuamba | chlorfenapyr<br>+ pyrethroid<br>ITN | 79,551 | 267,444 | 74,092 | 0.93 |
|  | Mandima | pyriproxyfen<br>+ pyrethroid<br>ITN | 44,614 | 198,625 | 24,865 | 0.56 |
| Mozambique<br>(West) | Guro | chlorfenapyr<br>+ pyrethroid<br>ITN | 33,543 | 112,753 | 89,367 | 2.66 |
|  | Tambara | chlorfenapyr<br>+ pyrethroid<br>ITN | 29,460 | 62,091 | 49,213 | 1.67 |
|  | Changara | PBO +<br>pyrethroid<br>ITN | 3,769 | 136,595 | 20,236 | 5.37 |
|  | Doa | PBO +<br>pyrethroid<br>ITN | 4,913 | 96,656 | 14,319 | 2.91 |
| Rwanda | Karongi | chlorfenapyr<br>+ pyrethroid<br>ITN | 124,212 | 418,146 | 187,391 | 1.51 |
|  | Ruhango | IRS +<br>standard ITN | 260,188 | 403,120 | 4,837,440 | 18.59 |

*All cost values are reported in 2020 USD. Incremental costs are relative to distribution of standard ITNs alone.*

Costs per case averted were less than 10 USD per case in all study areas. The highest cost per case averted involved the use of IRS rather than a dual-AI or PBO ITN but also resulted in the highest numbers of cases averted per person targeted.

### Sensitivity analyses

Uncertainty in costs per case averted was captured by Monte Carlo simulation from the effect models and of cost data points (Figure 7).

**Figure 7:**
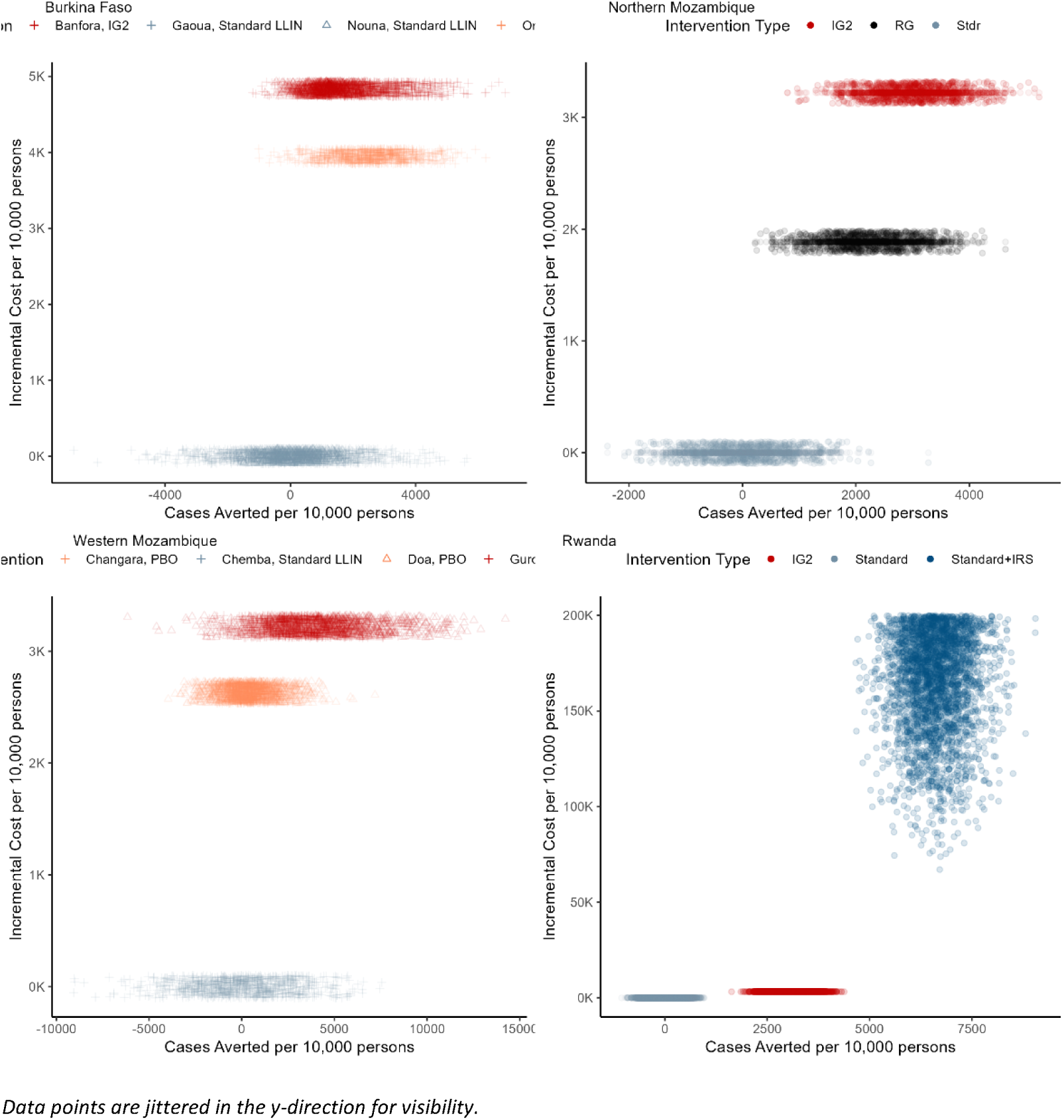
Cost-effectiveness results.

Cost-effectiveness data clouds show that variability in cost-effectiveness was driven by price differences and overlapping effect estimates in these studies. As cost of distribution was generally treated as identical in pairs of distribution simulations (cost of distribution was assumed to be variable but independent of ITN type for a specific simulation) cost differences in ITN programs are thus driven solely by prices of ITNs which were fixed at the prices described above. Cost-effectiveness data clouds were further analyzed by creating cost-effectiveness acceptability curves (Figure 8).

**Figure 8:**
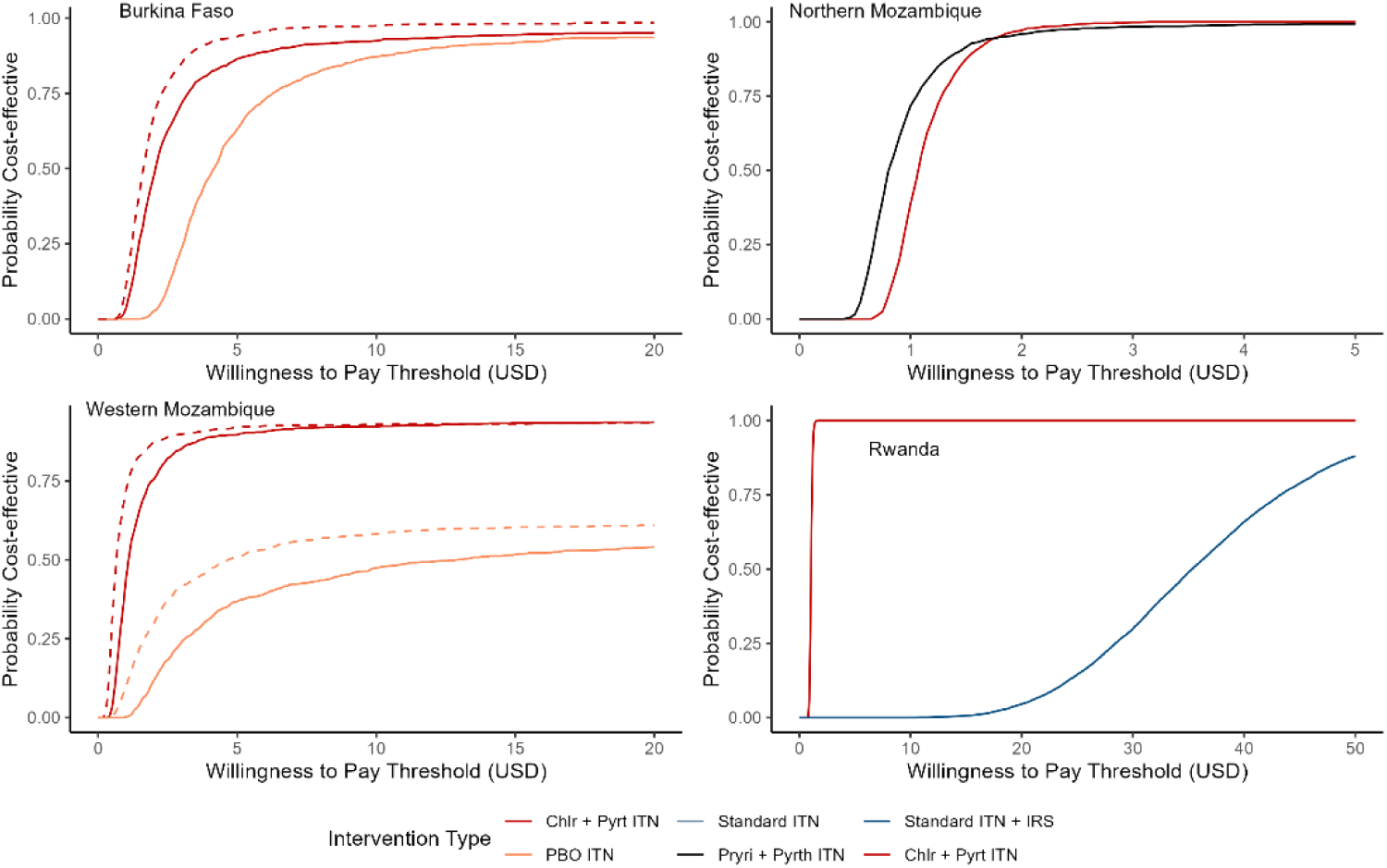
Cost-effectiveness acceptability curves for individual site level analysis.

A global meta-analysis was conducted for each ITN type (and IRS) to determine the expected relative cost-effectiveness across a number of settings of malaria transmission assuming that the relative effect size (incidence rate ratio) was independent of transmission setting.

Cost and interventions effects summarized across the studies were synthesized to provide a global analysis of estimated ICER per DALY averted for varied levels of baseline incidence. The results are shown in Figure 9.

**Figure 9:**
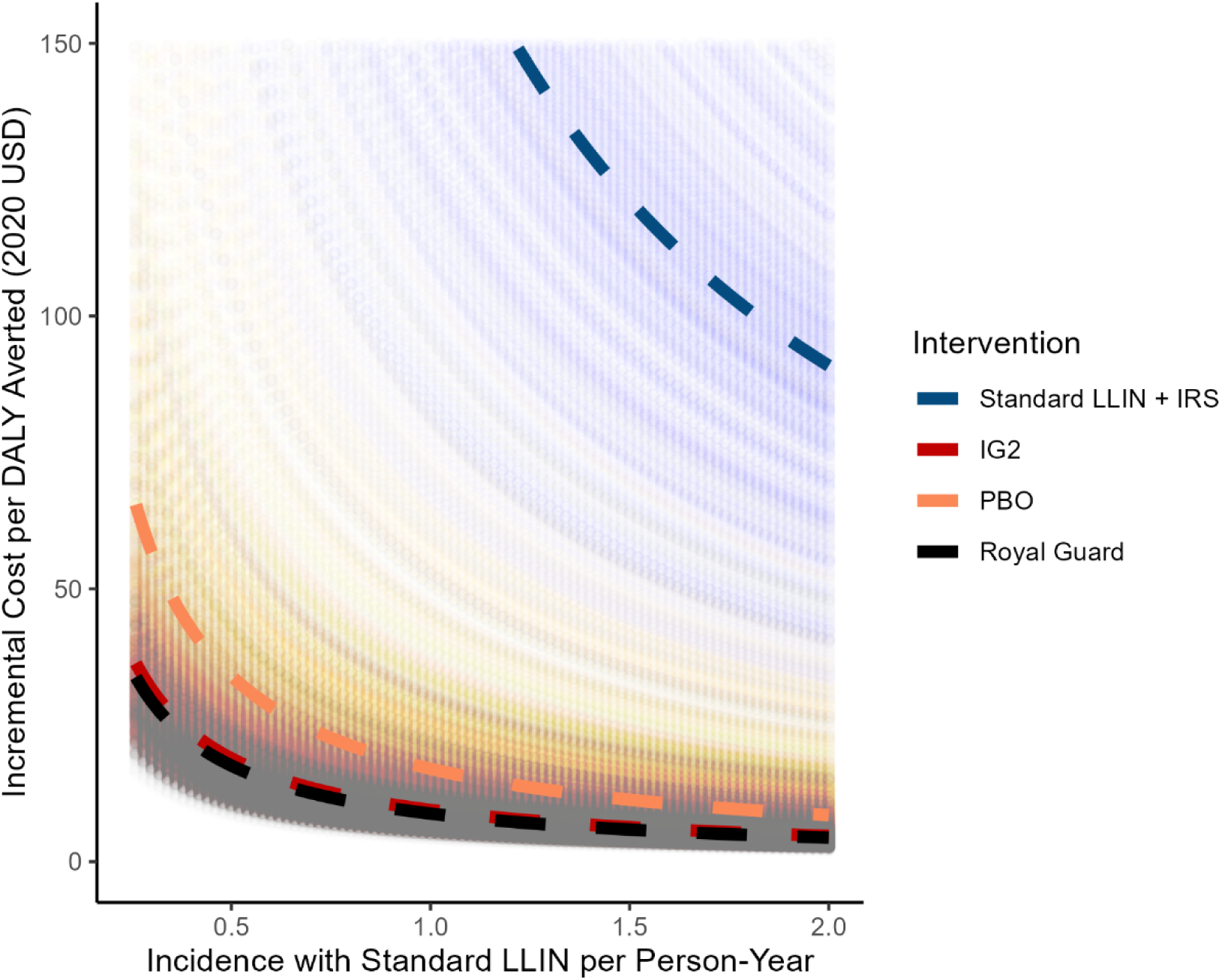
Global Sensitivity Analysis

High variability in effect estimates leads to significant overlap in ICER estimates between the three dual-AI or PBO ITN types, though chlorfenapyr + pyrethroid nets and pyriproxyfen + pyrethroid nets are clearly superior in cost-effectiveness to PBO + pyrethroid nets, and in all cases the incremental cost-effectiveness of using a dual-AI or PBO ITN type appears to be clearly lower than adding IRS to a standard ITN distribution. This remains true despite the effect of adding IRS being much larger than that of using dual-AI or PBO ITNs.

### Budget impact

Impact on budget was assessed in both gross and ITN cost approaches. Gross budget impact was not sensitive to underlying malaria incidence because the interventions are deployed to an entire population irrespective of the local malaria burden. Adding IRS is generally more expensive at more than 5 USD per person-year at risk, while all dual-AI or PBO ITNs added approximately 0.50 USD per person-year when compared to deployment of standard ITNs over a three-year time period. ITN costs (for all dual-AI or PBO ITN interventions) were generally cost saving at all but the lowest incidence levels when it was assumed that cost savings could be recouped linearly for every case which was averted. For the addition of IRS it was only cost saving on average when incidence was shown to be above approximately one case per person year (Figure 10).

**Figure 10:**
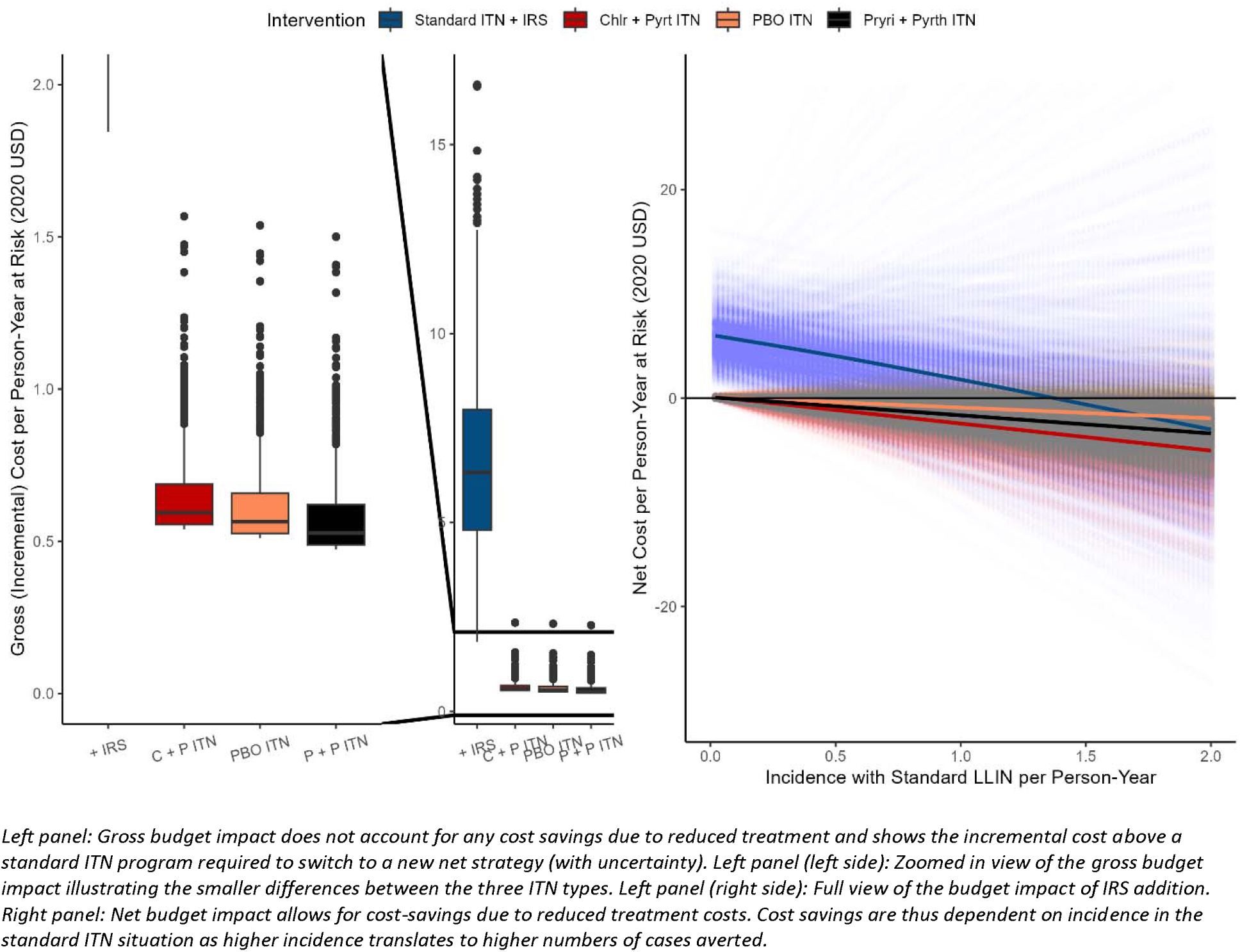
Budget Impact Analysis

Left panel: Gross budget impact does not account for any cost savings due to reduced treatment and shows the incremental cost above a standard ITN program required to switch to a new net strategy (with uncertainty). Left panel (left side): Zoomed in view of the gross budget impact illustrating the smaller differences between the three ITN types. Left panel (right side): Full view of the budget impact of IRS addition. Right panel: Net budget impact allows for cost-savings due to reduced treatment costs. Cost savings are thus dependent on incidence in the standard ITN situation as higher incidence translates to higher numbers of cases averted.

## DISCUSSION

The cost-effectiveness of dual-AI or PBO ITN products, namely chlorfenapyr + pyrethroid ITNs, pyriproxyfen + pyrethroid ITNs, and PBO + pyrethroid ITNs, as well as IRS + standard ITNs is of significant interest to malaria programs and global malaria funders. This study aimed to assess cost-effectiveness and budget impact of deploying these interventions to provide valuable insights for decision-makers working in resource-constrained settings.

One of the main strengths of this research lies in its comprehensive evaluation of multiple intervention strategies in real-world contexts. By comparing the cost-effectiveness of the available dual-AI or PBO ITNs and IRS combined with standard ITNs, we were able to capture the potential benefits and drawbacks of each approach. This multi-site, multi-intervention analysis allows us to consider a range of options for malaria control, catering to different operational contexts and constraints as well as to provide data that supports the benchmarking of operational decisions about malaria vector control throughout sub-Saharan Africa.

The findings of our study demonstrated that all four interventions would be generally considered cost-effective in reducing malaria transmission. The incremental cost per DALY averted was below the standard threshold of most sub-Saharan African countries’ GDP per capita, indicating the attractiveness of using a dual-AI or PBO ITN in most endemic settings. Chlorfenapyr + pyrethroid ITNs, which showed higher efficacy against pyrethroid-resistant mosquitoes, showed the highest impact in terms of DALYs averted of the three ITN types. Pyriproxyfen + pyrethroid ITNs and PBO + pyrethroid ITNs also yielded substantial health gains, albeit at a slightly higher cost per case averted. IRS combined with standard ITNs was the most effective intervention (although limited data was generated in this study from only one study location) and presented a cost-effective alternative for settings with larger budgets.

Moreover, the economic analysis considered not only the direct costs of the interventions but also the potential savings in healthcare expenditures due to reduced malaria morbidity and mortality. By incorporating these cost offsets, we were able to provide a sense of the potential budget impact of the interventions in both gross and net impact terms. This approach offers a more comprehensive perspective for decision-makers, taking into account the potential cost savings associated with effective malaria control.

However, it is important to acknowledge the limitations of this study. First, the cost-effectiveness estimates were based on data from observational studies which may be more at risk of some kinds of bias than controlled randomized studies. While we aimed to use the most reliable and up-to-date information on cost for ITNs and IRS, there is inherent uncertainty in such projections. Decision makers will need to reassess both the primary information on the cost of interventions and the likely effects in context to ensure that the results from this range of studies also translate meaningfully into their settings.

Another limitation is the lack of consideration for potential implementation challenges and operational constraints. While these data were derived from real-world implementation, further use of interventions may still not achieve the levels of coverage and compliance rate with the intervention strategies seen in these pilot settings. Factors such as insecticide resistance, cultural acceptability, and logistical barriers could impact the actual effectiveness and cost-effectiveness of these interventions.

In the context of previous research, our study builds upon the existing evidence on the cost-effectiveness of malaria control interventions. While previous studies have evaluated the economic efficiency of ITNs or IRS individually, our research contributes by comparing multiple dual-AI or PBO ITN products and evaluating their cost-effectiveness against with IRS + standard ITNs. This analysis provides decision-makers with a more nuanced understanding of the trade-offs and potential synergies between different interventions, enabling them to make informed choices based on their specific context and available resources.

Recent massive cuts in the funding for global malaria control due to the dismantling of USAID and PMI make information on the efficient use of resources in malaria control all the more valuable. While these cuts will undoubtedly have serious impacts on malaria mortality and morbidity this year and moving forward (3), identifying and deploying the most cost-efficient malaria control strategies available under the remaining budgets may help to minimize the impact of these actions.

## CONCLUSION

This study highlights the cost-effectiveness of chlorfenapyr + pyrethroid nets, pyriproxifen + pyrethroid nets, PBO + pyrethroid nets, and IRS combined with standard ITNs as malaria control strategies. The use of chlorfenapyr + pyrethroid nets appear most cost-effective across a wide range of settings, but other ITN-based strategies are also attractive and differences may be small. The addition of IRS to standard ITNs appears to be a highly effective but expensive strategy. These interventions have the potential to yield substantial health benefits while remaining economically viable in resource-constrained settings. Uncertainty around the relative ranking of the three ITN types remains substantial, and the need for decisions to account for local variation in effectiveness and cost for local implementation remains. In the current context understanding which interventions and combinations are expected to be most cost-effective and efficient is critical.

## Data Availability

All data produced in the present study are available upon reasonable request to the authors

